# Development of a digitally-obtainable 10-year all-cause mortality risk score based on data from 497,712 UK Biobank participants

**DOI:** 10.1101/2021.06.23.21259387

**Authors:** Michele Colombo, Nikola Dolezalova, Aleksa Despotovic, Angus B. Reed, Davide Morelli, Mert Aral, David Plans

**Affiliations:** Huma Therapeutics Ltd, London, United Kingdom; Faculty of Medicine, University of Belgrade, Serbia; Department of Engineering Science, Institute of Biomedical Engineering, University of Oxford, Oxford, United Kingdom; Department of Experimental Psychology, University of Oxford, Oxford, United Kingdom; INDEX Group, Department of Science, Innovation, Technology, and Entrepreneurship, University of Exeter, United Kingdom

**Keywords:** Mortality, prediction model, machine learning, survival analysis, digital health

## Abstract

**Background:** All-cause mortality (ACM) scores are a useful tool for identifying individuals with decreased life expectancy. An interpretable score consisting of smartphone-obtainable variables could allow for long-term management of individual health and support the next generation of healthcare monitoring and preventative practices. The aim of this study was to develop a 10-year ACM risk score using the UK Biobank dataset, using only digitally-obtainable variables.

**Methods:** The models were developed using the full UK Biobank cohort comprising nearly 500,000 individuals. We extracted 399 features from the dataset and, through a data-driven feature selection process with subsequent clinical review, identified 34 features for the final model. As part of the study, we compared two survival analysis approaches: Cox proportional hazards model and DeepSurv, a deep learning-based survival analysis algorithm.

**Results:** Before feature selection, Cox performed similarly to DeepSurv, achieving a c-index of 0.771 (95% CI 0.770–0.772) and 0.774 (95% CI 0.772–0.775) on the test dataset, respectively. Using the selected 34 features, the c-index of Cox decreased slightly to 0.770 (95% CI 0.769–0.770) and DeepSurv to 0.758 (95% CI 0.755–0.762). The models show excellent calibration at 10 years.

**Conclusions:** This study improves on a previous smartphone-compatible score, C-Score, by incorporating non-modifiable factors in addition to variables which can be actively modified to reduce risk. This score is comprehensive, easily interpretable and actionable, and as such, could provide a powerful tool for preventative healthcare.

## Introduction

The rapid increase in life expectancy and decrease in birth rates in many countries around the world in recent decades has brought about a change in demographic landscape ^1,2^. Populations are ageing, conferring increased healthcare expenditure due to the higher number of morbidities in the elderly and the average higher cost per morbidity in this demographic ^3^. Being able to identify individuals who have decreased life expectancy has important implications for policy and clinical practice, as well as for the individuals themselves, particularly if they are supported in identifying any pathways to reduce this risk, such as by changing certain lifestyle factors. Prognostic models of death from any cause (‘all-cause mortality’, ACM) over a specified time period have been a helpful tool for evaluation of overall health status.

The National Institute for Health Care and Excellence (NICE) reviewed 41 existing tools for mortality predictions in 2016. It recommended that, owing to ubiquitous, shared limitations, further research should be undertaken to develop reliable tools for use in clinical practice ^4^. Many of these predictive models were developed using cohorts of older individuals (>65 years) with a prediction horizon between one and five years ^5–8^. The UK Biobank (UKB) ^9^, a cohort study of ∼500,000 UK participants aged 38-73, provides a unique opportunity to study risk factors for a broader age range over a longer time period.

Implementing an ACM score in a smartphone application would maximise access to tools that could support individuals’ long-term health management. Such a score should be easily interpretable, actionable, and visibly dynamic to incentivise sustained lifestyle changes. Indeed, modifiable risk factors such as tobacco use, activity, and diet have been shown to be strongly associated with mortality ^10–12^ and subsequently used in other risk models ^13^. Our previous effort to build a risk score within a smartphone application, named C-Score ^14^, incorporated heart rate, sleep duration, waist-to-height ratio, number of cigarettes per day, alcohol intake, reaction time, and self-rated health for predictions of 10-year ACM. This score deliberately included only modifiable predictors, resulting in a concordance index (c-index) of 0.66.

Here, we aim to build from this proof of concept and expand potential predictors to medical history, family history, sociodemographic and environmental factors, physical activity, mental health, and diet; many of which are known predictors of mortality ^7,15,16^. All variables available for most UKB participants will be used in the initial set, following the exclusion of those that are not easily acquired by smartphone (via user input or passive recording) or are country-specific. Contrary to previous studies, we aim to use an entirely data-driven approach to select the most significant predictors from this initial set of variables, with a clinical review of the final predictor selection. Our modelling approach comprises traditional Cox proportional hazards modelling alongside a machine learning approach to survival analysis, the Cox proportional hazards deep neural network (DeepSurv) ^17^.

This study aims to develop a data-driven prognostic model for 10-year ACM using the UK Biobank dataset that can be implemented in a smartphone setting to support user engagement with their health.

## Methods

### Study Population

Data comes from the UKB ^9^, approved under UKB application number 55668. UKB participants were recruited for a prospective cohort study from the general population between 2006 and 2010. Data up to the 30th September 2020 update were used, which we further consider as the end of the follow-up period.

### Input Features

We selected 77 fields based on literature review and clinical plausibility, ensuring that the information could be collected on a smartphone and applied to different geographies. This initial set included basic demographics (age, sex, education level), anthropometrics (body measurements, weight, BMI), biometrics (heart rate), alcohol and smoking habits, sleep habits, self-rated health, medical and family history, physical activity habits, dietary habits, UV exposure and protection, and environmental variables (air pollution, proximity to roads).

### Preprocessing

ACM outcome was defined as death from any cause during the follow-up period as per UKB field 40000. Additional insights were obtained analysing the underlying causes of death, field 40001. The length of follow-up was defined as the period between assessment date and either date of death or the end date of the study.

Main data transformations were: mean-imputation of missing values; merging groups of highly specific fields into a summary field (e.g. average weekly alcohol consumption was derived from a sum of consumption of different drink types); merging sex-specific fields (e.g. male-only and female-only fields for various medications); or deriving ratios of original features (e.g. waist-to-height ratio). Lastly, all categorical information was one-hot encoded, followed by excluding categories occurring less than 0.1%. Processing steps are summarised in ***Supplementary Table 1***.

### Experimental Settings

The dataset was split into training (75%) and test (25%) sets; the latter was used only for the final model’s validation.

Two survival analysis approaches were tested, the Cox Proportional Hazard (CPH) model ^18^ and its deep learning variant, DeepSurv ^17^, which exploits artificial neural networks to model the relationship between prognostic factors and survival time. In the first instance, we used CPH to minimise the number of features without significant performance degradation. Both CPH and DeepSurv were then trained and evaluated using the resulting set of features.

### CPH Model and Feature Selection

As CPH models are semi-parametric, the model’s selection phase practically reduces to feature selection only.

Using the *lifelines* package^19^ an initial model was obtained by adjusting for age only. A baseline model with all the features was then trained and a stepwise variable selection process employed to remove features which do not have significant impact on performance. A set of six features (nine following one-hot encoding of self-rated health) was manually fixed within the model to extend the previously developed C-Score ^14^.

We trained a univariate model for each feature during forward selection, keeping only those with *p*-value <0.10. A model was trained with all the remaining candidate variables during backward selection and its performance assessed using 5-fold cross-validation. Models excluding features in decreasing *p*-value order were then tested and if performance did not significantly degrade, the feature was eliminated. The process was continued until all variables were tested for removal. Features were initially tested in chunks of decreasing size in order to accelerate the process.

The final step of feature selection involved manual review in which features were eliminated where they were deemed clinically insignificant and where there was minimal performance contribution among the initially fixed features.

### DeepSurv

DeepSurv models ^17^, in contrast to the CPH model, require extensive hyperparameter optimisation. The focus, at first, was finding the best hyper-parameterisation for the replete baseline model to assess whether the problem involved non-linear components that the CPH model would not capture. A separate set of optimal hyperparameters was defined for the final reduced model using the same procedure. Since results suggested no significant improvement could be achieved by using DeepSurv on the baseline input space, no further experiments for features selection using DeepSurv were performed.

Models were trained employing an extension of the deep learning library *PyTorch* ^20,21^. Hyperparameter space was explored through a Tree-Structured Parzen Estimator (TPE) ^22^, as provided by the *Optuna* library ^23^. Each model was tested employing three-fold cross-validation. Feed-forward neural networks with up to three hidden layers were tested, details of methods and search space are provided in ***Supplementary Table 2***.

### Statistical analysis

Statistical analysis of baseline characteristics and train and test datasets were performed using Python *tableone* library ^24^. The discrimination metric for all models was the concordance index (c-index), while the Integrated Calibration Index (ICI), implemented in the lifelines library^19^, was used to evaluate calibration at the 10-year timepoint. Confidence intervals (CIs) were obtained using percentile bootstrap resampling with 50 resampling rounds.

## Results

### Population characteristics

The entire UKB cohort was used in this study. After excluding participants with missing data, the dataset contained 497,712 participants. There were 29,615 (5.96%) participants who died during follow-up (***Figure 1a***). There were no statistical differences between train and test datasets among the features included in the final model (***Supplementary Table 3***).

**Figure 1:**
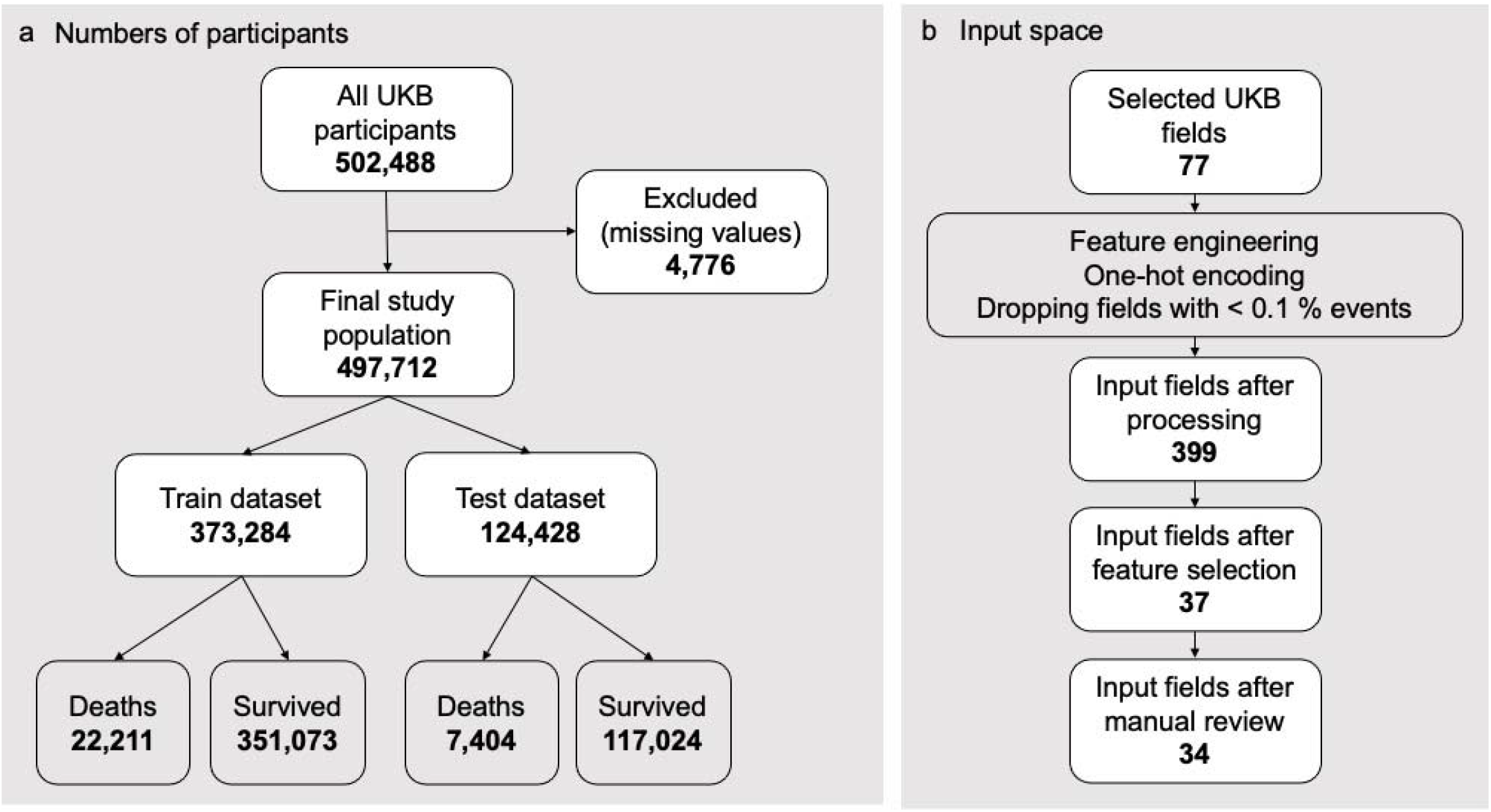
Flow diagram of participants and input variables in the study. **(a)** Participant numbers used in the study, including breakdown of the recorded death outcomes in the train and test datasets. **(b)** Size of the input space before and after processing and after feature selection.

The analysis of mortality causes in the studied cohort is summarised in ***Supplementary Table 4*** and revealed that 53.3% of the deaths resulted from cancers (most commonly lung, breast, and pancreas cancers) and 20.3% from diseases of the cardiovascular system (particularly chronic ischaemic heart disease, myocardial infarction, and stroke). The remainder of the top-5 are diseases of the respiratory (7.3%), nervous (4.9%), and digestive system (3.8%). All other causes each contributed <3% of the total deaths.

The demographic analysis of the cohort is presented in ***Supplementary Table 5***, both in the overall sample and separated by outcome. Among the participants, 54.4% were women, with a median age of 58 at recruitment, and predominantly white (>94%). The median follow-up time was 11.6 years (IQR 10.87–12.33).

### Feature selection and CPH model

Model performance is reported in ***Table 2***. The CPH model comprising only age obtained 0.690 c-index on the training dataset and 0.694 on the test dataset. The model trained with all 399 input features led to a c-index of 0.779 (95% CI 0.778–0.779) on the training dataset and 0.771 (95% CI 0.770–0.772) in the test dataset.

**Table 2:** CPH models results reported for different sets of input features. Shown are concordance indices obtained during training and internal validation on the test dataset, along with 95 % bootstrap confidence intervals.

| Features Set | Number of input features | Train C-Index | Test C-Index |
| --- | --- | --- | --- |
| Age | 1 | 0.6900 [0.6900–0.6900] | 0.6940 [0.6940–0.6940] |
| All Features | 399 | 0.7786 [0.7783–0.7790] | 0.7712 [0.7702–0.7719] |
| Step-Wise Selected | 37 | 0.7724 [0.7721–0.7727] | 0.7697 [0.7694–0.7700] |
| Manually Reviewed | 34 | 0.7722 [0.7719–0.7725] | 0.7695 [0.7691–0.7697] |

***Supplementary Table 1*** outlines the features selected according to the stepwise variable selection procedure. Numbers of input features in the individual steps of the feature selection process are also summarised in ***Figure 1b***. 80 features were removed from the candidate set without any measurable degradation of performance following forward selection. Following backward elimination, 37 features were selected. These features were further subjected to manual review, excluding initially ‘fixed’ features with negligible impact (sleep duration and cigarettes-per-day) or those with problematic clinical explanation (experienced headaches in the past month being a protective feature), resulting in 34 features. The performance after manual review remained equivalent: 0.772 on the training dataset and 0.770 on the test dataset. The contribution of individual features to the overall performance is shown in ***Supplementary Figure 1***, while the plot of coefficients for individual features is presented in ***Figure 2*** (detailed results in ***Supplementary Table 6***).

**Figure 2:**
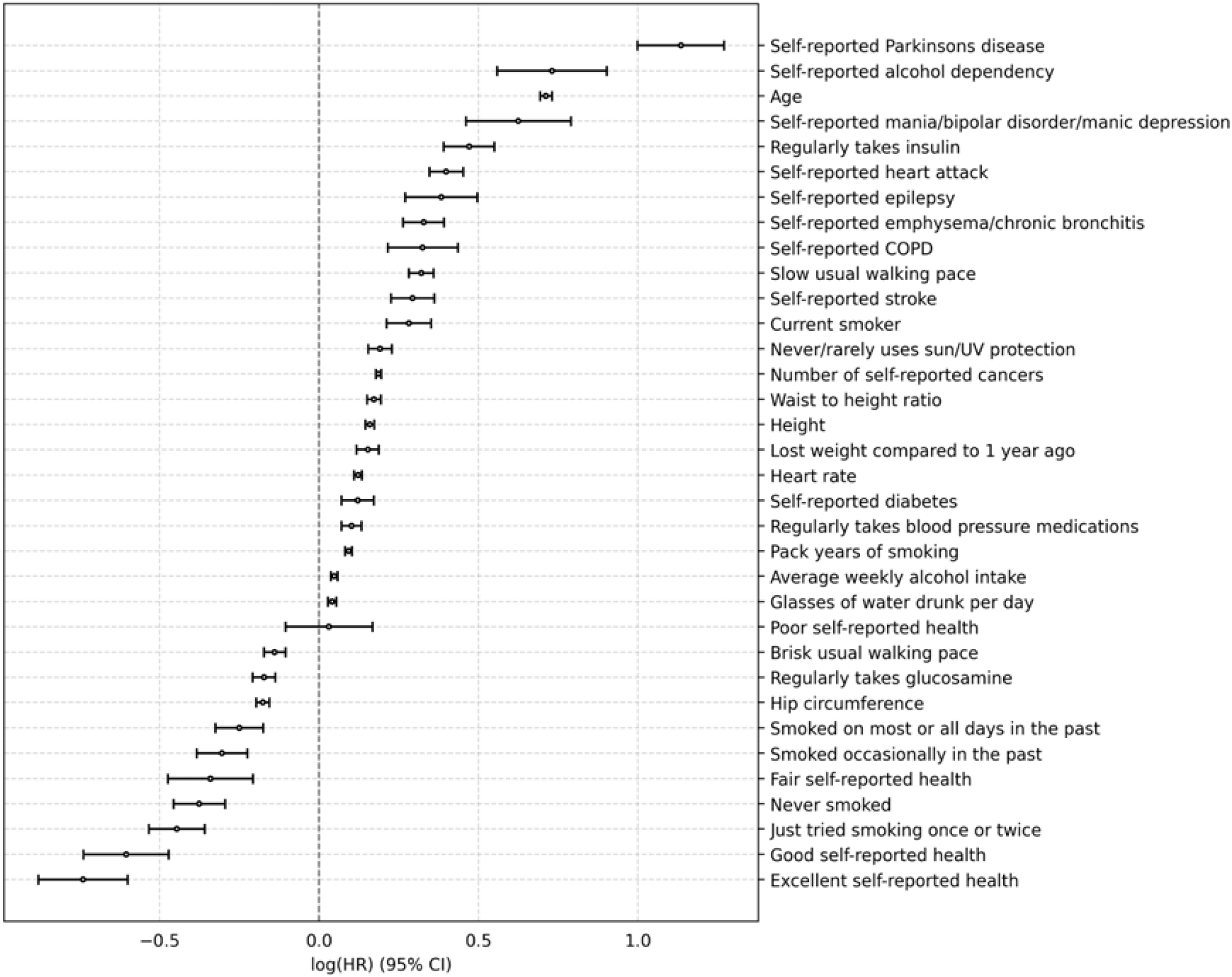
Plot of Cox Proportional Hazards model coefficients. Points show log(HR) ± 95% CI. HR = hazard ratio, CI = confidence interval.

While the baseline model slightly overestimated the predicted risk (ICI 0.10%), the final model showed excellent calibration (ICI 0.03%, ***Supplementary Figure 2***). The mean observed 10-year risk in the cohort was 4.79% (95% CI 4.75–4.82), while the 10-year risk predicted by the final model was 4.82% (95% CI 4.78–4.85).

### DeepSurv

Optimal hyperparameters for baseline (399 features) and final (34 features) model were selected using three-fold cross-validation. Performance comparable to the CPH model was obtained in fewer than 50 iterations of the TPE algorithm (***Supplementary Figure 3***). Subsequently, only negligible performance improvement was achieved. Hence, we limited the number of trials to 200 to avoid potential overfitting.

The resulting hyperparameters for the baseline model led to a c-index of 0.774 (95% CI 0.772–0.775; trial 85) in the test dataset. For the final model with 34 features, the best performance was 0.758 (95% CI 0.755–0.762; trial 181). There was minimal difference between performance on the training and test datasets for both models, indicating no overfitting (***Table 2***).

**Table 2:** Best baseline and reduced models selected within first 200 trials. Median concordance indices with 95% bootstrap confidence intervals shown.

|  | Trial # | Hyper-Parameters | Train C-Index | Test C-Index |
| --- | --- | --- | --- | --- |
| Baseline model<br>(399 features) | 85/200 | Activation: SELU<br>Batch Normalization: Yes<br>Dropout: 0.1311<br>Weight Decay: 0.0402<br>Learning Rate: 0.010119<br>Optimizer: SGD<br>Momentum: 0.3650<br>Hidden Layers Shape: 128x128 | 0.7795<br>[0.7785–0.7806] | 0.7736<br>[0.7724–0.7746] |
| Final model<br>(34 features) | 181/200 | Activation: ReLU<br>Batch Normalization: Yes<br>Dropout: 0.1585<br>Weight Decay: 12.8300<br>Learning Rate: 0.000550<br>Optimizer: Adam<br>Hidden Layers Shape: 256 | 0.7620<br>[0.7571–0.7652] | 0.7584<br>[0.7546–0.7618] |

## Discussion

By virtue of the UKB’s comprehensive and diverse data, coupled with a long follow-up period, we were able to create a 10-year ACM CPH model with excellent predictive capability. Age and age-related conditions such as Parkinson’s disease, which is known to contribute to ACM ^25^, were predictably identified as both having high importance to the model alongside high hazard ratios (HR). Additionally, a number of pre-existing conditions, including cardiovascular (stroke and myocardial infarction), respiratory (COPD, emphysema, and bronchitis), diabetes, cancer, and psychiatric and neurological disorders, significantly contribute to ACM in our model. All retained pre-existing conditions are known to affect life expectancy ^26–30^. The majority of these conditions are non-communicable diseases, which are largely preventable through appropriate modifications in lifestyle and behavioural aspects of health ^31^, as well as early medical intervention.

Besides pre-existing conditions, the features with the highest HR in our model — alcohol dependency, slow usual walking pace, active smoking, higher waist-to-height ratio, and increased resting heart rate — have all previously been shown to contribute to ACM ^32^. These features point to the fundamental aspects of one’s health and their relationship with ACM, specifically physical activity, nutrition, alcohol intake, and smoking status ^32^. Interestingly, never or rarely using UV protection was another lifestyle factor that is significantly associated with increased risk for ACM in our model. The relationship between UV exposure and development of skin cancers has been established in the literature ^33^, but the exact long-term effects of sunscreen protection are yet to be fully understood ^34^.

Contrastingly, the bulk of protective factors are common knowledge — brisk walking pace, positive self-reported health, and a never-smoker status or history of smoking cessation. Again, this points to the preventable aspect of disease occurrence, and emphasising again the well known benefit of smoking cessation even after years of smoking ^35^. Lastly, regular glucosamine use was identified as protective in our model. Often used for treatment of joint pain, glucosamine’s beneficial effect on ACM has been established in literature by reducing one’s risk of developing several age-related diseases ^36^.

In addition to the CPH model, we tested the deep learning approach to survival analysis, DeepSurv. This model achieved comparable performance for the baseline model with all 399 features but slightly underperformed CPH for the final model. The lack of significant improvement when implementing deep learning is not uncommon with ACM, as was shown in ^17^, seemingly as minimal contribution of non-linear associations between factors; thus DeepSurv’s ability to take advantage of non-linear relationships has not been exploited in this setting. Additionally, there is limited interpretability of the individual feature contributions in black-box models such as DeepSurv, making them less suitable for clinical translation.

Our model significantly improves on the previously published smartphone-compatible algorithm, C-Score, achieving a c-index of 0.77 vs. 0.66, respectively ^14^. Among other studies using UKB, Ganna and Ingelsson (2015) built a CPH model for the prediction of 5-year ACM, achieving a c-index of 0.80 for men and 0.79 for women ^8^. Separately, Weng *et al*. employed both a traditional statistical approach (c-index 0.75) and machine learning (0.78–0.79) to train models for prediction of 10-year premature ACM ^37^. Unlike these studies, we employed survival analysis in both traditional statistical and machine learning modeling which allowed us to account for length of survival rather than binary outcome at a single time point. Compared to our results, these models contain notable differences in the final features, likely due to different methodological approaches to feature selection. Our selection process allowed us to create a geographically-agnostic model (e.g. absence of UK-specific ‘Townsend deprivation Index’), which requires at the minimum only an internet connection to complete, while still maintaining good predictive capability.

The value in such a model is two-fold: first, if used on an individual level, accessible ACM models can form the backbone of behaviour-change programmes by presenting the user with interactable, dynamic health forecasts based on their lifestyle choices; second, if used on a regional or population level, such models could be used to inform local funding initiatives targeted to the most prevalent risk factors within their sub-population.

The primary limitation of this study concerns the UKB dataset. First, the majority of the UKB population is of White ethnicity (94%), which can lead to poor replicability when implemented across other ethnic groups. Second, the cohort’s age range is restricted to 37-73 years, which may impart a similar impact on generalisability. Third, the UKB population is considered to be healthier and wealthier than the general population ^38^. These limitations mean external validation is needed to solidify its applicability both in the UK and across other populations.

We have developed a 10-year ACM model with very good predictive capability that can be readily accessible through smartphones by the general population. A focus on factors that are modifiable either by an individual or at a population level further supports the needed shift towards preventative healthcare and promotes longevity. Future studies on more diverse samples should be carried out to enable its widespread use.

## Data Availability

Data cannot be shared publicly owing to the violation of patient privacy and the absence of informed consent for data sharing.

## Acknowledgements

The authors would like to acknowledge Adam Cunningham for his contribution during the preparation of this manuscript.

## Supplementary information

**Supplementary Figure 1:**
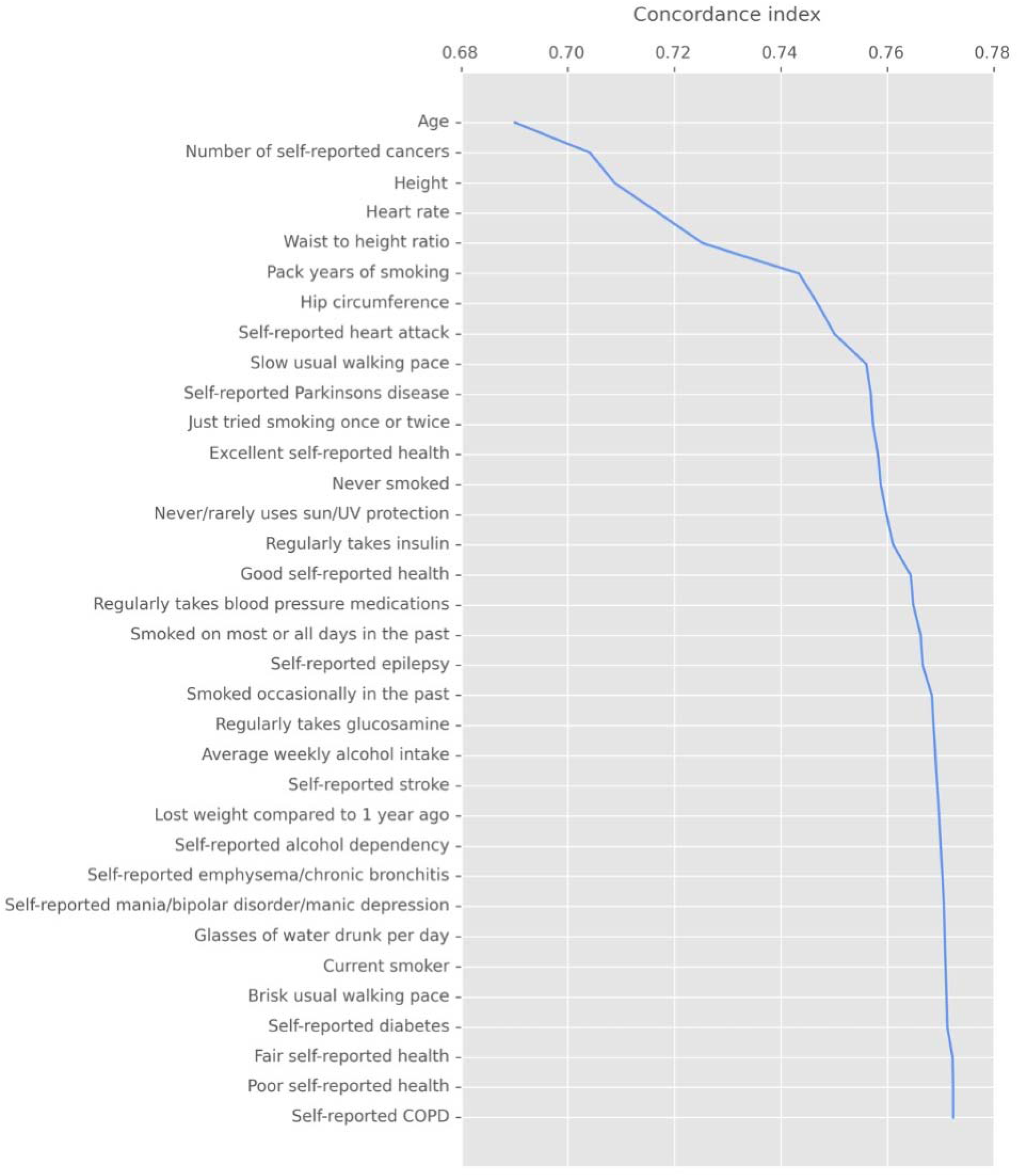
Contribution of features to the final model concordance index. Features were added stepwise from the top, in the order of permutation importance (i.e. age, being the most important feature, was added first, self-reported COPD last to complete the feature set and achieve the final concordance index).

**Supplementary Figure 2:**
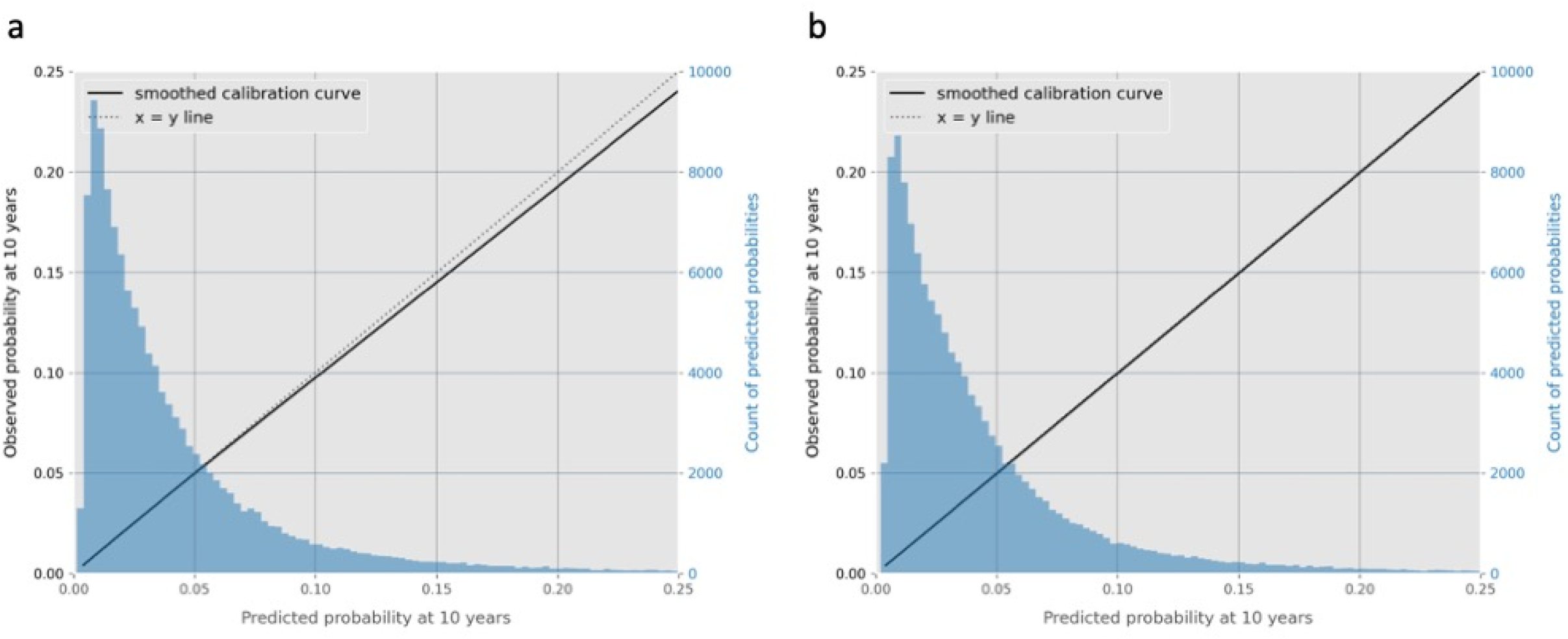
Model calibration at 10 years. Results from the baseline (a) and final (b) CPH models evaluated on the test dataset are shown. Smoothed calibration curve is shown in solid line. Histogram of the predicted probabilities of incident death at 10 years for the participants in the test dataset are shown in blue.

**Supplementary Figure 3:**
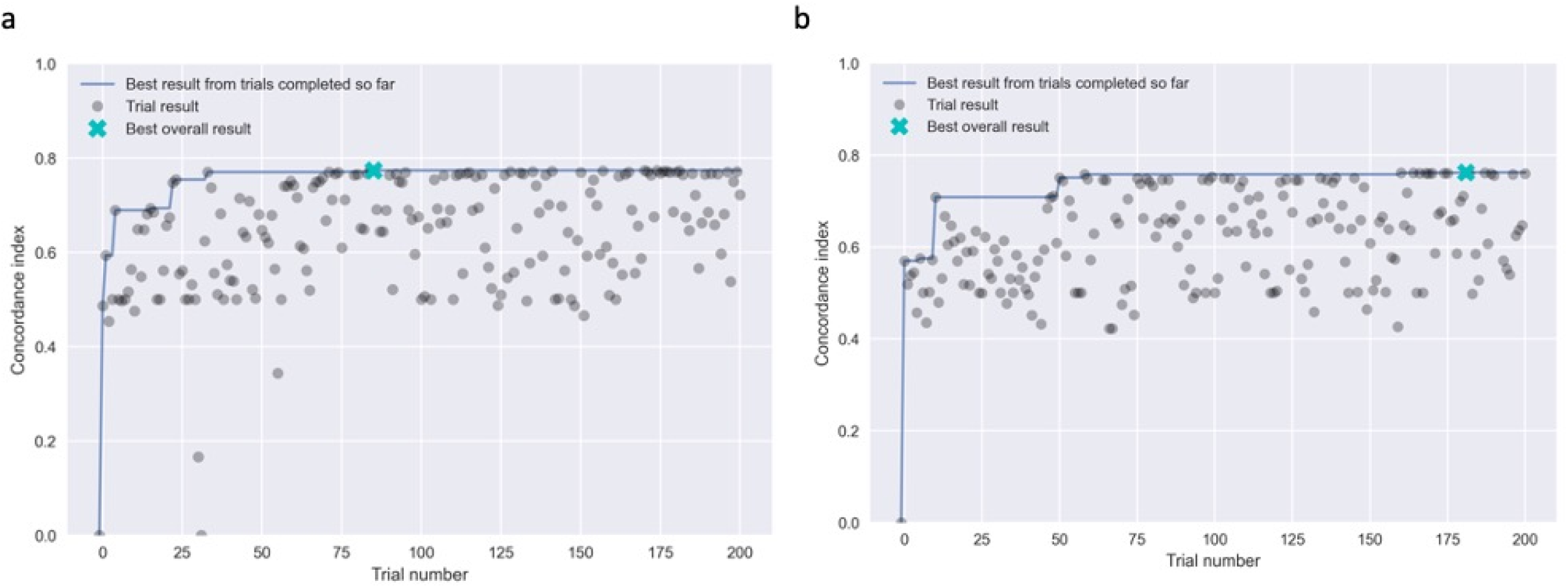
**Performance achieved in the 200 trials of TPE hyperparameter search** for the baseline (a) and final (b) model. Trials with the best overall results are indicated with a cyan cross.

**Supplementary Table 1:**
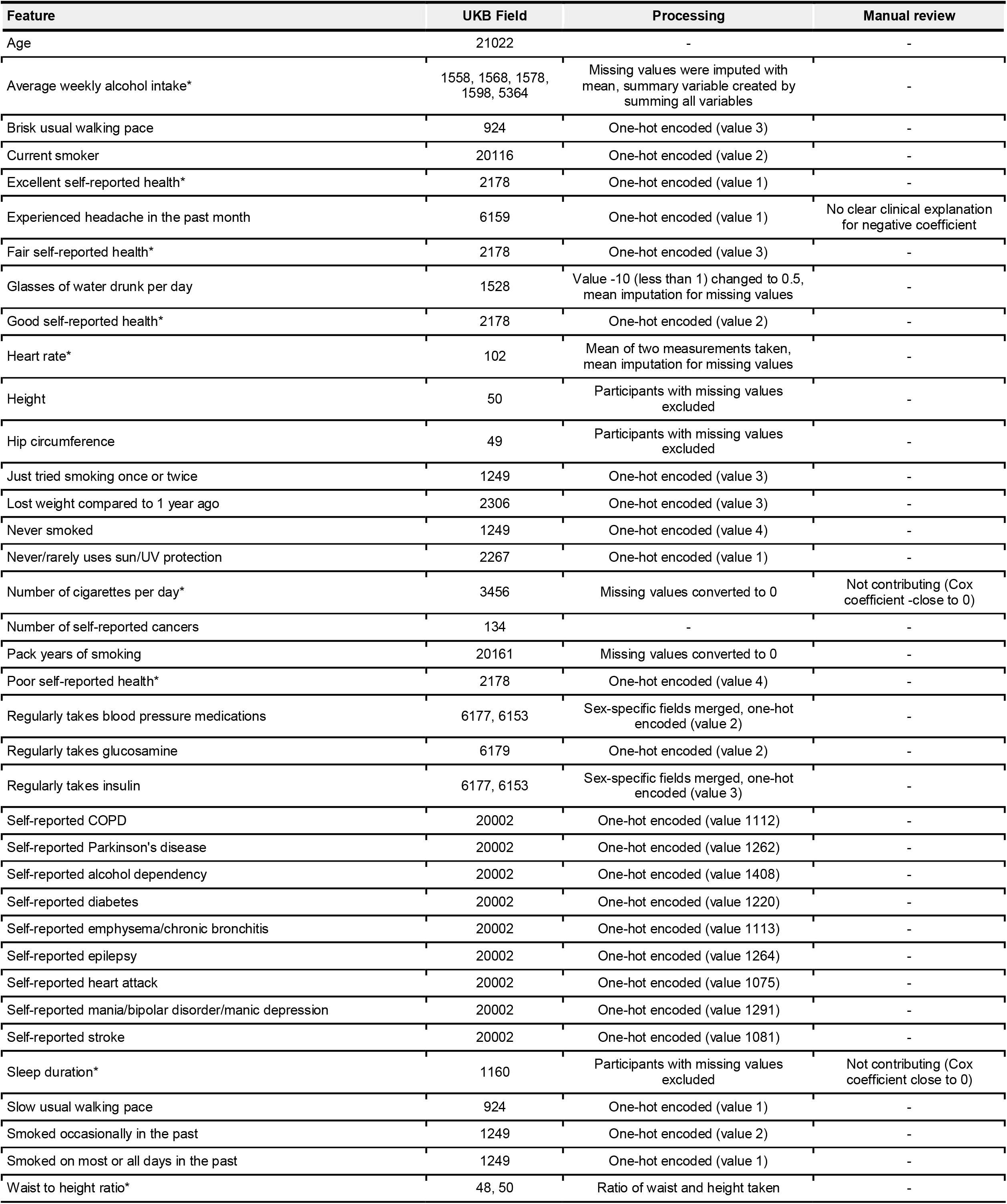
List of features selected during the data-driven feature selection process. Source UK Biobank field along with any data preprocessing methods are shown. Features fixed during the feature selection process are marked with an asterisk. Three features were removed during manual review and the reasons are summarised in the last column.

**Supplementary Table 2:**
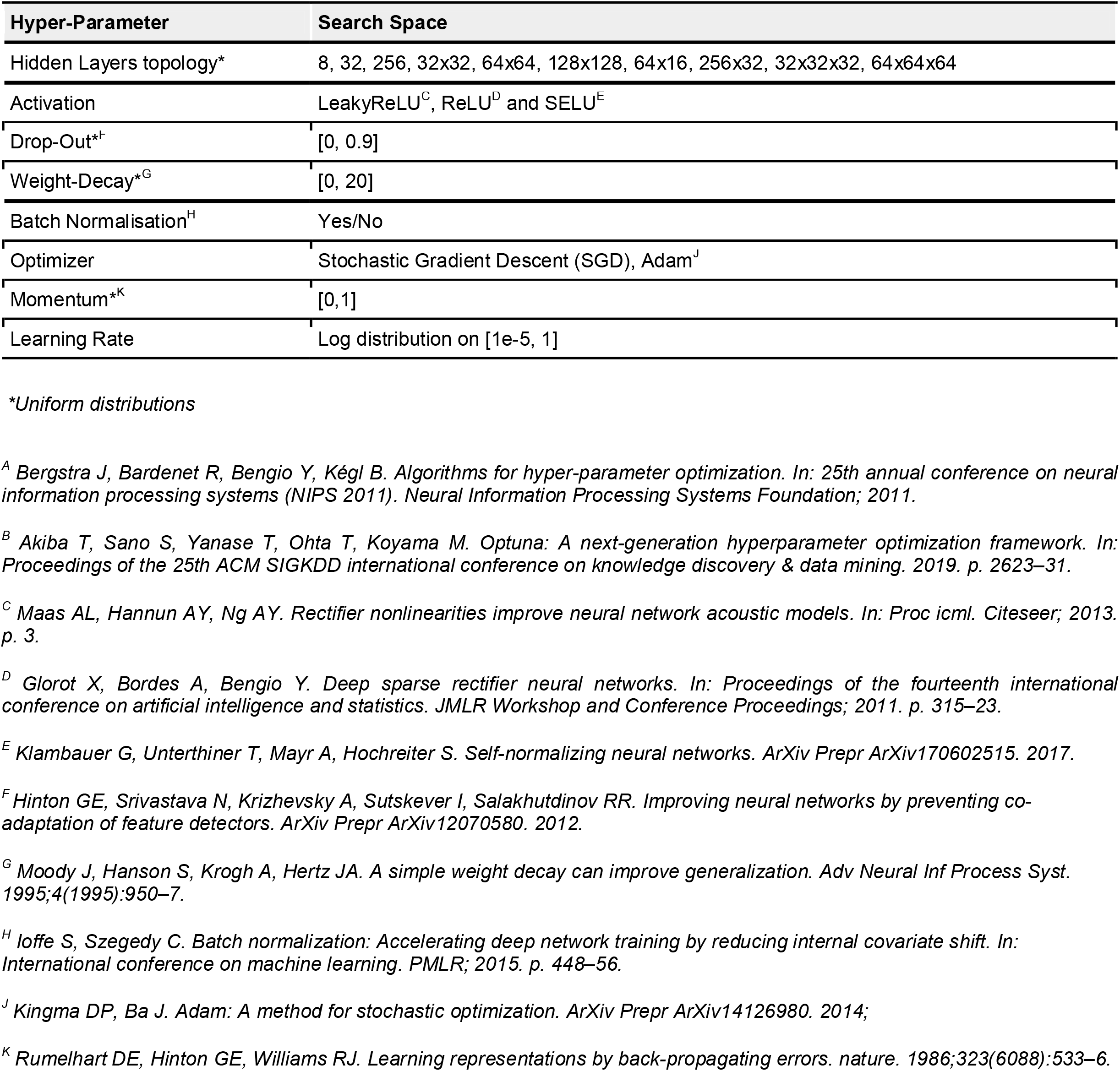
DeepSurv hyperparameter search space. Tree-Structured Parzen Estimator algorithm^A^ from the Optuna library^B^ was used to find the optimal set of parameters within the search space.

**Supplementary Table 3:** Statistical comparison of the train and test datasets. Features selected into the final model are shown in alphabetical order. Last column shows p-value after comparing the incident death group with the no-death group. Comparisons were performed using the Chi-squared test for categories and Kruskal-Wallis test for continuous variables.

|  | Overall | Train dataset | Test dataset | p-value (adjusted) |
| --- | --- | --- | --- | --- |
| n | 497712 | 373284 | 124428 |  |
| Death (outcome), n (%) | 29615 (5.95) | 22211 (5.95) | 7404 (5.95) | 1.000 |
| Follow-up time, median [Q1, Q3] | 11.60<br>[10.88,12.32] | 11.60<br>[10.88,12.32] | 11.60<br>[10.88,12.32] | 1.000 |
| Age, median [Q1,Q3] | 58.00<br>[50.00,63.00] | 58.00<br>[50.00,63.00] | 58.00<br>[50.00,63.00] | 0.298 |
| Average weekly alcohol intake, median [Q1,Q3] | 11.48 [6.04,12.00] | 11.48 [6.04,12.00] | 11.48 [6.04,12.00] | 1.000 |
| Brisk usual walking pace, n (%) | 193135 (38.80) | 144893 (38.82) | 48242 (38.77) | 1.000 |
| Current smoker, n (%) | 52401 (10.53) | 39320 (10.53) | 13081 (10.51) | 1.000 |
| Excellent self-reported health, n (%) | 81486 (16.37) | 61223 (16.40) | 20263 (16.28) | 1.000 |
| Fair self-reported health, n (%) | 104277 (20.95) | 78037 (20.91) | 26240 (21.09) | 1.000 |
| Glasses of water drunk per day, median [Q1,Q3] | 2.00 [1.00,4.00] | 2.00 [1.00,4.00] | 2.00 [1.00,4.00] | 1.000 |
| Good self-reported health, n (%) | 287268 (57.72) | 215412 (57.71) | 71856 (57.75) | 1.000 |
| Heart rate, median [Q1,Q3] | 69.00<br>[62.00,75.50] | 69.00<br>[62.00,75.50] | 69.00<br>[62.00,75.50] | 1.000 |
| Height, median [Q1,Q3] | 168.00<br>[162.00,175.00] | 168.00<br>[161.50,175.00] | 168.00<br>[162.00,175.00] | 1.000 |
| Hip circumference, median [Q1,Q3] | 102.00<br>[97.00,108.00] | 102.00<br>[97.00,108.00] | 102.00<br>[97.00,108.00] | 0.278 |
| Just tried smoking once or twice, n (%) | 72575 (14.58) | 54525 (14.61) | 18050 (14.51) | 1.000 |
| Lost weight compared to 1 year ago, n (%) | 75218 (15.11) | 56147 (15.04) | 19071 (15.33) | 0.542 |
| Never smoked, n (%) | 199306 (40.04) | 149424 (40.03) | 49882 (40.09) | 1.000 |
| Never/rarely uses sun/UV protection, n (%) | 50190 (10.08) | 37600 (10.07) | 12590 (10.12) | 1.000 |
| Number of self-reported cancers, median [Q1,Q3] | 0.00 [0.00,0.00] | 0.00 [0.00,0.00] | 0.00 [0.00,0.00] | 1.000 |
| Pack years of smoking, median [Q1,Q3] | 0.00 [0.00,7.12] | 0.00 [0.00,7.00] | 0.00 [0.00,7.50] | 1.000 |
| Poor self-reported health, n (%) | 22209 (4.46) | 16745 (4.49) | 5464 (4.39) | 1.000 |
| Regularly takes blood pressure medications, n (%) | 102974 (20.69) | 77097 (20.65) | 25877 (20.80) | 1.000 |
| Regularly takes glucosamine, n (%) | 93995 (18.89) | 70339 (18.84) | 23656 (19.01) | 1.000 |
| Regularly takes insulin, n (%) | 5482 (1.10) | 4108 (1.10) | 1374 (1.10) | 1.000 |
| Self-reported COPD, n (%) | 1644 (0.33) | 1219 (0.33) | 425 (0.34) | 1.000 |
| Self-reported Parkinson's disease, n (%) | 840 (0.17) | 628 (0.17) | 212 (0.17) | 1.000 |
| Self-reported alcohol dependency, n (%) | 737 (0.15) | 542 (0.15) | 195 (0.16) | 1.000 |
| Self-reported diabetes, n (%) | 21387 (4.30) | 16055 (4.30) | 5332 (4.29) | 1.000 |
| Self-reported emphysema/chronic bronchitis, n (%) | 6744 (1.36) | 5087 (1.36) | 1657 (1.33) | 1.000 |
| Self-reported epilepsy, n (%) | 3975 (0.80) | 2977 (0.80) | 998 (0.80) | 1.000 |
| Self-reported heart attack, n (%) | 11352 (2.28) | 8473 (2.27) | 2879 (2.31) | 1.000 |
| Self-reported mania/bipolar disorder/manic depression, n (%) | 1400 (0.28) | 1030 (0.28) | 370 (0.30) | 1.000 |
| Self-reported stroke, n (%) | 6546 (1.32) | 4905 (1.31) | 1641 (1.32) | 1.000 |
| Slow usual walking pace, n (%) | 40144 (8.07) | 30131 (8.07) | 10013 (8.05) | 1.000 |
| Smoked occasionally in the past, n (%) | 64998 (13.06) | 48888 (13.10) | 16110 (12.95) | 1.000 |
| Smoked on most or all days in the past, n (%) | 120108 (24.13) | 89981 (24.11) | 30127 (24.21) | 1.000 |
| Waist to height ratio, median [Q1,Q3] | 0.53 [0.48,0.58] | 0.53 [0.48,0.58] | 0.53 [0.48,0.58] | 0.149 |

**Supplementary Table 4:** Analysis of the most common causes of death in the dataset. ICD10 codes belonging in each group are listed in the second column. Number and percentage of participants who died during follow-up are shown, along with 3 most common ICD10 codes in each group.

|  | ICD10 codes | Number (%) of participants | Top 3 ICD10 codes |
| --- | --- | --- | --- |
| **Any cause** | **any** | **29615 (100 %)** | **C34 Malignant neoplasm of bronchus and lung**<br>**I25 Chronic ischaemic heart disease**<br>**I21 Acute myocardial infarction** |
| Neoplasms | C00–C97<br>D10–D48 | 15790<br>(53.32 %) | C34 Malignant neoplasm of bronchus and lung<br>C50 Malignant neoplasm of breast<br>C25 Malignant neoplasm of pancreas |
| Diseases of the circulatory system | I05–I89 | 6085<br>(20.55 %) | I25 Chronic ischaemic heart disease<br>I21 Acute myocardial infarction<br>I64 Stroke, not specified as haemorrhage or infarction |
| Diseases of the respiratory system | J09–J99 | 2157<br>(7.28 %) | J44 Other chronic obstructive pulmonary disease<br>J84 Other interstitial pulmonary diseases<br>J18 Pneumonia, organism unspecified |
| Diseases of the nervous system | G00–G99 | 1442<br>(4.87 %) | G30 Alzheimer's disease<br>G12 Spinal muscular atrophy and related syndromes<br>G20 Parkinson's disease |
| Diseases of the digestive system | K20–K93 | 1125<br>(3.80 %) | K70 Alcoholic liver disease<br>K55 Vascular disorders of intestine<br>K74 Fibrosis and cirrhosis of liver |
| External causes of morbidity and mortality | V01–V97<br>W00–W99<br>X00–X99<br>Y10–Y89 | 861<br>(2.91 %) | X70 Intentional self-harm by hanging, strangulation and suffocation<br>W19 Unspecified fall<br>Y83 Surgical operation and other surgical procedures as the cause of abnormal reaction of the patient, or of later complication, without mention of misadventure at the time of the procedure |
| Mental and behavioural disorders | F00–F89 | 568<br>(1.92 %) | F03 Unspecified dementia<br>F01 Vascular dementia<br>F10 Mental and behavioural disorders due to use of alcohol |
| Other unknown or unspecified causes | R00–R99<br>U00–U49 | 460<br>(1.55 %) | U07 Emergency use of U07<br>R99 Other ill-defined and unspecified causes of mortality<br>U50 Level of care administered to neonates |
| Endocrine, nutritional and metabolic diseases | E00–E90 | 337<br>(1.14 %) | E11 Non-insulin-dependent diabetes mellitus<br>E14 Unspecified diabetes mellitus<br>E85 Amyloidosis |
| Certain infectious and parasitic diseases | A00–A99<br>B00–B99 | 264<br>(0.89 %) | A41 Other septicaemia<br>A81 Atypical virus infections of central nervous system<br>A09 Diarrhoea and gastro-enteritis of presumed infectious origin |
| Diseases of the genitourinary system | N00–N98 | 189<br>(0.64 %) | N39 Other disorders of urinary system<br>N18 Chronic renal failure<br>N17 Acute renal failure |
| Diseases of the musculoskeletal system and connective tissue | M00–M90 | 168<br>(0.57 %) | M06 Other rheumatoid arthritis<br>M35 Other systemic involvement of connective tissue<br>M34 Systemic sclerosis |
| Diseases of the blood and blood-forming organs and certain disorders involving the immune mechanism | D55–D89 | 69<br>(0.23 %) | D86 Sarcoidosis<br>D70 Agranulocytosis<br>D61 Other aplastic anaemias |
| Diseases of the skin and subcutaneous tissue | L00–L99 | 49<br>(0.17 %) | L03 Cellulitis<br>L97 Ulcer of lower limb, not elsewhere classified<br>L08 Other local infections of skin and subcutaneous tissue |
| Congenital malformations, deformations and chromosomal abnormalities | Q00–Q99 | 48<br>(0.16 %) | Q87 Other specified congenital malformation syndromes affecting multiple systems<br>Q24 Other congenital malformations of heart<br>Q61 Cystic kidney disease |
| Diseases of the ear and mastoid process | H65–H75 | 2<br>(0.01 %) | H70 Mastoiditis and related conditions |
| Pregnancy, childbirth and the puerperium | O00–O99 | 1<br>(0.00 %) | O30 Multiple gestation |

**Supplementary Table 5:** Summary of demographic characteristics of the studied cohort grouped by the outcomes. Features selected into the final model are shown in alphabetical order. Last column shows p-value after comparing the incident death group with the no-death group. Comparisons were performed using the Chi-squared test for categories and Kruskal-Wallis test for continuous variables.

|  | Overall | No death during follow-up | Death during follow-up | p-value (adjusted) |
| --- | --- | --- | --- | --- |
| n | 497712 | 468097 | 29615 |  |
| Age, median [Q1,Q3] | 58.00<br>[50.00,63.00] | 57.00<br>[50.00,63.00] | 63.00<br>[58.00,67.00] | <0.001 |
| Average weekly alcohol intake, median [Q1,Q3] | 11.48<br>[6.04,12.00] | 11.48<br>[6.04,12.00] | 11.48<br>[8.00,13.04] | <0.001 |
| Brisk usual walking pace, n (%) | 193135 (38.80) | 185802 (39.69) | 7333 (24.76) | <0.001 |
| Current smoker, n (%) | 52401 (10.53) | 46615 (9.96) | 5786 (19.54) | <0.001 |
| Excellent self-reported health, n (%) | 81486 (16.37) | 78779 (16.83) | 2707 (9.14) | <0.001 |
| Fair self-reported health, n (%) | 104277 (20.95) | 95291 (20.36) | 8986 (30.34) | <0.001 |
| Glasses of water drunk per day, median [Q1,Q3] | 2.00 [1.00,4.00] | 2.00 [1.00,4.00] | 2.00 [1.00,4.00] | <0.001 |
| Good self-reported health, n (%) | 287268 (57.72) | 273596 (58.45) | 13672 (46.17) | <0.001 |
| Heart rate, median [Q1,Q3] | 69.00<br>[62.00,75.50] | 69.00<br>[62.00,75.00] | 69.50<br>[63.50,78.50] | <0.001 |
| Height, median [Q1,Q3] | 168.00<br>[162.00,175.00] | 168.00<br>[161.20,175.00] | 169.00<br>[162.00,176.00] | <0.001 |
| Hip circumference, median [Q1,Q3] | 102.00<br>[97.00,108.00] | 102.00<br>[97.00,108.00] | 103.00<br>[98.00,109.00] | <0.001 |
| Just tried smoking once or twice, n (%) | 72575 (14.58) | 69887 (14.93) | 2688 (9.08) | <0.001 |
| Lost weight compared to 1 year ago, n (%) | 75218 (15.11) | 69944 (14.94) | 5274 (17.81) | <0.001 |
| Never smoked, n (%) | 199306 (40.04) | 190784 (40.76) | 8522 (28.78) | <0.001 |
| Never/rarely uses sun/UV protection, n (%) | 50190 (10.08) | 45305 (9.68) | 4885 (16.50) | <0.001 |
| Number of self-reported cancers, median [Q1,Q3] | 0.00 [0.00,0.00] | 0.00 [0.00,0.00] | 0.00 [0.00,0.00] | <0.001 |
| Pack years of smoking, median [Q1,Q3] | 0.00 [0.00,7.12] | 0.00 [0.00,6.00] | 0.00 [0.00,27.30] | <0.001 |
| Poor self-reported health, n (%) | 22209 (4.46) | 18246 (3.90) | 3963 (13.38) | <0.001 |
| Regularly takes blood pressure medications, n (%) | 102974 (20.69) | 92102 (19.68) | 10872 (36.71) | <0.001 |
| Regularly takes glucosamine, n (%) | 93995 (18.89) | 89125 (19.04) | 4870 (16.44) | <0.001 |
| Regularly takes insulin, n (%) | 5482 (1.10) | 4374 (0.93) | 1108 (3.74) | <0.001 |
| Self-reported COPD, n (%) | 1644 (0.33) | 1194 (0.26) | 450 (1.52) | <0.001 |
| Self-reported Parkinson's disease, n (%) | 840 (0.17) | 541 (0.12) | 299 (1.01) | <0.001 |
| Self-reported alcohol dependency, n (%) | 737 (0.15) | 562 (0.12) | 175 (0.59) | <0.001 |
| Self-reported diabetes, n (%) | 21387 (4.30) | 18143 (3.88) | 3244 (10.95) | <0.001 |
| Self-reported emphysema/chronic bronchitis, n (%) | 6744 (1.36) | 5353 (1.14) | 1391 (4.70) | <0.001 |
| Self-reported epilepsy, n (%) | 3975 (0.80) | 3551 (0.76) | 424 (1.43) | <0.001 |
| Self-reported heart attack, n (%) | 11352 (2.28) | 9065 (1.94) | 2287 (7.72) | <0.001 |
| Self-reported mania/bipolar disorder/manic depression, n (%) | 1400 (0.28) | 1213 (0.26) | 187 (0.63) | <0.001 |
| Self-reported stroke, n (%) | 6546 (1.32) | 5314 (1.14) | 1232 (4.16) | <0.001 |
| Slow usual walking pace, n (%) | 40144 (8.07) | 33927 (7.25) | 6217 (20.99) | <0.001 |
| Smoked occasionally in the past, n (%) | 64998 (13.06) | 61624 (13.16) | 3374 (11.39) | <0.001 |
| Smoked on most or all days in the past, n (%) | 120108 (24.13) | 110119 (23.52) | 9989 (33.73) | <0.001 |
| Waist to height ratio, median [Q1,Q3] | 0.53 [0.48,0.58] | 0.53 [0.48,0.58] | 0.56 [0.51,0.61] | <0.001 |

**Supplementary Table 6:**
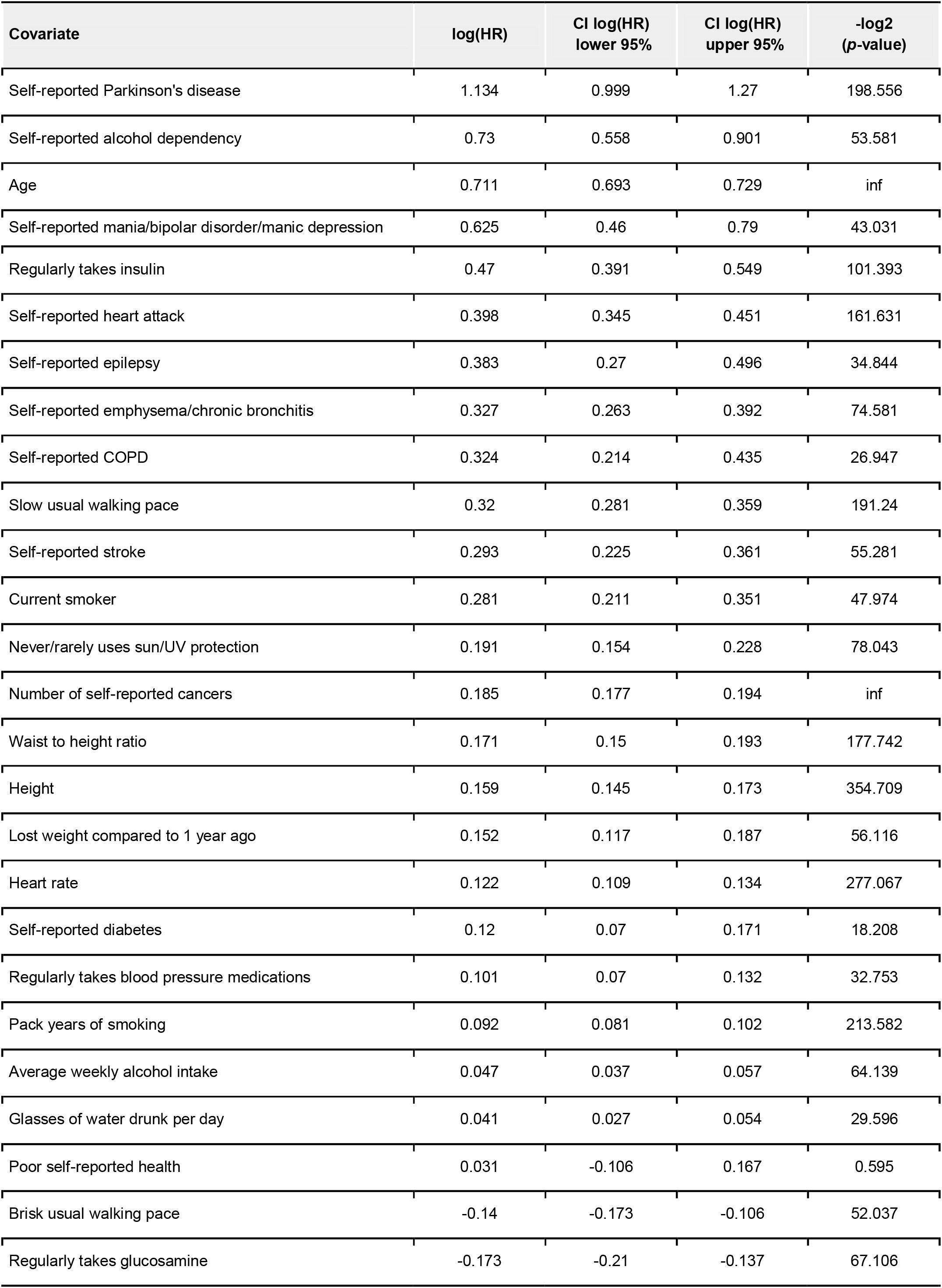
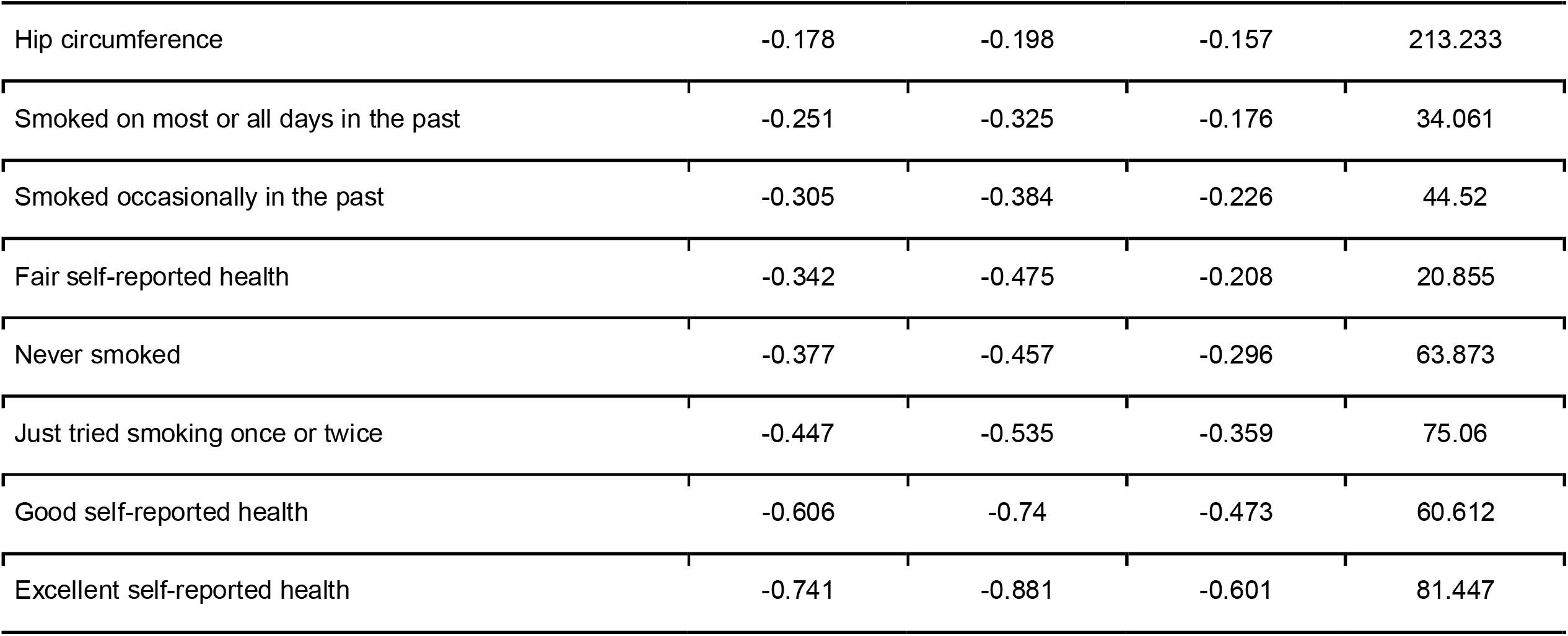
Summary of the final Cox Proportional Hazards model. The table displays coefficients = log(HR) with 95% confidence intervals and −log2(p-value). All columns were statistically significant (where p < 0.05 and null hypothesis states that the coefficient is equal to 0) except “Poor self-reported health” where the p-value was 0.662.

## Notes

**Conflict of Interest:** MC, ND, AD, ABR, DM, MA, and DP are employees of Huma Therapeutics Ltd.

### Competing Interest Statement

MC, ND, AD, ABR, DM, MA, and DP are employees of Huma Therapeutics Ltd.

### Funding Statement

This work was funded by Huma Therapeutics Ltd

### Author Declarations

Data comes from the UKB, approved under UKB application number 55668

